# Structure of anxiety associated with the COVID-19 pandemic in the Russian-speaking sample: results from on-line survey

**DOI:** 10.1101/2020.04.28.20074302

**Authors:** M.Yu. Sorokin, E.D. Kasyanov, G.V. Rukavishnikov, O.V. Makarevich, N.G. Neznanov, N.B. Lutova, G.E. Mazo

**Affiliations:** V.M. Bekhterev National Medical Research Center for Psychiatry and Neurology, Saint-Petersburg, Russia; I.P. Pavlov First Saint-Petersburg State Medical University, Saint-Petersburg, Russia

**Keywords:** coronavirus infection, pandemic, COVID-19, mental health, anxiety, affective disorders, stigmatization

## Abstract

The COVID-19 pandemic imposed not only serious threats to the physical health of the population, but also provoked a wide range of psychological problems.

**Objective:** To identify the most vulnerable populations during the epidemic period (including individuals with affective disorders) who are most in need of psychological and / or psychiatric help.

**Material and methods:** On-line survey of 1957 Russian-speaking respondents over 18 years old from March 30 to April 5, 2020. The level of anxiety distress was verified with the psychological stress scale (PSM-25). Stigmatization of individuals experiencing respiratory symptoms was assessed with modified devaluation / discrimination questionnaire (PDD; Cronbach’s α = 0.707).

**Results:** 99.8% of respondents had variable concerns associated with COVID-19. Their mean scores of psychological stress were increased to moderate levels (104.9 ± 34.4 points), and the stigmatization scores exceeded the value of the whole sample median (19.5 ± 3.4; Me = 19). 35% of respondents had concerns about COVID-19 associated with anxiety distress (Cohen’s d = 0.16–0.39): these were “risk of isolation” and “possible lack of medication for daily use”. The most prone to concerns were respondents’ groups with affective disorders, young people (≤20 years old), unemployed, single, those without higher education and women.

**Conclusions:** Large sub-cohorts of the Russian-speaking sample need correction of anxiety distress associated with the COVID-19 pandemic. The implementation of such measures should be targeted and oriented in terms of coverage and content to identified vulnerable social groups.

---

First cases of a novel coronavirus infection, caused by the SARS-CoV-2 coronavirus, (COVID-19 from COrona VIrus Disease 2019) were detected in November 2019 [1]. This infection spread quickly in Wuhan (the capital of the Chinese province of Hubei) and then throughout whole China, later spreading to other countries including the Russian Federation (RF) and leading to a global health emergency [2]. As early as March 11, 2020, due to the high prevalence of COVID-19 cases, the World Health Organization (WHO) recognized the current situation as a pandemic [3]. In the Russian Federation, the first patients with COVID-19 were identified on January 31, 2020, and in early April, more than 50,000 Russians had confirmed diagnoses [4,5].

The COVID-19 pandemic imposed serious threats to physical health and individual’s lives. Moreover, the risk of coronavirus infection caused a wide range of psychological problems, such as panic disorder, anxiety, and depression, among the population of countries with a high spread of viral infection [6]. Since March 2020, many governments around the world have introduced special quarantine measures to limit the spread of the virus and minimize the burden on medical services. Persons over 65, those with concomitant illnesses and pregnant women were asked to isolate themselves from direct contact with people for at least 12 weeks, and those suspected of carrying coronavirus were instructed to remain in their homes and isolate themselves and everyone living with them, for at least 14 days [7].

Thus, current situation includes a number of factors that significantly affect the mental health of the population at the same time:

1. An unprecedented, potentially life-threatening situation of indefinite duration;
2. Large-scale quarantine measures in all major cities, which essentially limit residents to their homes;
3. The indefinite incubation period of a viral infection and its possible asymptomatic transmission;
4. Reports of a lack of medical resources;
5. Unstable information background with an overabundance of controversial data;
6. Uncertainty associated with the possible consequences of coronavirus infection COVID-19 on the economic situation in the country.

According to Chinese researchers, the COVID – 19 coronavirus infection pandemic provoked a parallel epidemic of anxiety and depressive reactions [8, 9]. Moreover, certain groups of the population may be more vulnerable to psychological stress associated with this disease. This is especially true for individuals with affective disorders who are more susceptible to the COVID-19 pandemic emotional responses, which could manifest in mental symptoms relapses or worsening. It is due to this individual’s high sensitivity to stress compared to the general population, and also due to limitations of the scheduled psychiatric outpatient appointments. Furthermore, in addition to rising stress levels among the population, stigma and discrimination against certain populations’ increase [10], even in the absence of evidence of increased morbidity risks among discriminated groups.

**The aim** of this study was to identify the most vulnerable groups of the population during the epidemic period (including among individuals with affective disorders) who are most in need of psychological and / or psychiatric care.

## MATERIAL AND METHODS

### Setting and sample

The study was performed in accordance with the World Medical Association Declaration of Helsinki (2013). Approval was obtained from the local IRB at V.M. Bekhterev National Medical Research Center for Psychiatry and Neurology. All participants gave their consent to the processing of personal data before enrollment.

The research data were obtained from an on-line survey conducted from March 30 to April 5, 2020. Participants were asked to fill out the questionnaire at online platform Google Forms, which on average required about 15 minutes. The questionnaire was distributed in social networks, on the websites of public organizations and thematic communities (see Acknowledgements).

The inclusion criteria were: the age of participants over 18 years; ability to read and understand text in Russian; consent of personal data processing, the fact of which was considered in completion of all proposed survey forms. Non-inclusion criteria were the absence of data for any individual items of the questionnaire.

The questionnaire included questions about socio-demographic data of respondents, presence / absence of affective disorders (Major depressive disorder, Bipolar affective disorders, Generalized anxiety disorder, Dysthymia, Cyclothymia), information about somatic pathology. All data was obtained from self-report of study participants.

Participants were suggested to mark out any number of 10 items from the questionnaire describing various types of concerns associated with the COVID-19 pandemic. Furthermore, participants were asked to choose any number of 6 items describing behavioral patterns of infection prevention which were used by them (the full questionnaire is presented in Attachment). Also, respondents could describe the frequency of their information seeking requests about pandemic during last week. The request frequency was ranked by 8 points in the range from “never” to “hourly”. Psychological stress scale (PSM-25) was used to assess anxiety distress [11]. Stigmatization of individuals with signs of respiratory symptoms (cough, coryza, sneezing) was evaluated by modified version of popular questionnaire for perceived stigma evaluation, the section of devaluation/discrimination (PDD) [12]. The levels of agreement with questionnaire statements were evaluated by 4-point Likert scale. The higher total scores corresponded with more severe stigma intensity.

### Statistical analysis

Analysis was performed with Statistical Package for Social Science (SPSS), version 16.0 for Windows. Descriptive statistics were used. Normality test distribution was performed by using skewness and kurtosis values. Nominal scale dispersion was analyzed with Pearson’s χ^2^, Mann-Whitney U-criterion was used for ordinal scales differences. Size effects by Cohen’s d and Cramer’s V criteria were calculated for groups which had differences with statistical significance p≤0.05. Spearman’s correlation coefficient was calculated. Assessment of the internal consistency of the original stigma questionnaire was performed by using Cronbach’s α.

## RESULTS

The final poll included 2117 records. All questionnaires were filled during the first week of recommended self-isolation in Russia (from March 30 to April 5). Data of 160 respondents was excluded from analysis due to age criterion. Thus, statistical analysis was performed for 1957 respondents.

### Demographics

Majority of participants were women - 1649 respondents (84,3%). The mean age of all participants was 31±12 years. The sample was mostly from the populations of Federal cities - Saint Petersburg (21,1%), Moscow (16,8%), and also from all other Russian Federal Districts (57,6%), neighboring and foreign countries (4,5%).

### Social characteristics

About half of all the respondents had the college degree (55,3%). 25,6% participants reported incomplete higher education. Majority of respondents were employed in private (23,6%) and state (32,2%) organizations. Medical specialists were 10,3% of the sample. 22% participants had no regular employment. Unmarried/single were 51,8% of the sample. 26,9% were officially married, 12,4% lived in civil marriage.

### Concomitant disease

Respondents mentioned the presence of concomitant somatic pathology in 55,8% of cases. 29,5% participants confirmed that they had diagnoses of affective disorders. Most were Major depressive and Bipolar affective disorders (19,8%), less frequently - Anxiety disorders (6,0%) and Cyclothymia/Dysthymia (3,7%).

#### Characteristics of participants’ psychological and behavioral responses

The correlation analysis of the entire data set showed that adaptation to new living conditions during the spread of COVID-19 was a multi-level process with a complex structure of interrelated factors. The higher number of strategies taken to prevent coronavirus self-infection (on average 4 - Me; q25 = 3, q75 = 4) and the more frequent search for information about the epidemic (on average twice a day: Me = 6, q25 = 5, q75 = 7) predictably correlated in participants. They also were associated with an intensification of the psychological reactions of respondents to the pandemic. So, the number of anxious concerns and fears about COVID-19, the level of psychological stress associated with them, as well as the tendency of participants to stigmatize people with respiratory symptoms increased. Almost all of the above-mentioned items were also sensitive to the socio-demographic parameters of the research sample (table 1).

**Table 1.** Correlation of socio-demographic characteristics with the psychological and behavioral responses regarding COVID-19 (Spearman’s rho; N=1957; *p≤0.05, **p≤0.01, ***p≤0.001)

|  |  |  |  |  |  |
| --- | --- | --- | --- | --- | --- |
|  | Information search frequency | Themes of concerns | Preventive measures | Stress (PSM-25) | Stigmatization |
| Age | 0,06*** | -0,23*** |  | -0,38*** | 0,06** |
| Education level | 0,08*** | -0,15*** |  | -0,22*** |  |
| Information search frequency about COVID-19 | 1,000 | - | - | - | - |
| Number of concern themes about COVID-19 | 0,22*** | 1,000 | - | - | - |
| Number of measures to prevent COVID-19 self-infection | 0,17*** | 0,30*** | 1,000 | - | - |
| Psychological stress (PSM-25) | 0,14*** | 0,28*** | 0,05* | 1,000 | - |
| Stigmatization | 0,10*** | 0,12*** | 0,12*** |  | 1,000 |

Among the study participants, 99.8% reported at least 2 concern themes associated with the coronavirus, with the most common average number of 5 concern themes (Me=5; q25=4, q75=6, the typology is shown in Table 2). The prevalence of concerns was associated with a psychological stress indicator (PSM-25) reaching moderate levels (104.9±34.4 scores (M±S.D)) for the whole sample. A qualitative analysis of the interaction between individual concern themes associated with COVID-19 and the levels of psychological stress and stigmatization-discrimination of individuals with respiratory symptoms showed multidirectional effects of different apprehensions.

The presence of anxiety about the risk to the lives and health of relatives was not associated with a statistically significant increase in average stress levels or a significant increase in stigma. It was probably determined by the prevalence of this theme int majority of respondents. At the same time, a clinically significant effect of increasing psychological stress (weak in magnitude) was associated with the presence of 2 of the rarest of the 10 concern themes - “possible lack of medication for daily intake” and “risk of isolation” (table 2). In total, 688 individuals (35% of the sample) reported having at least one of them.

The average total score of the questionnaire on the stigmatization of individuals with respiratory symptoms was 19.5±3.4 (M±S.D.), with a median score of 19 and sufficient internal consistency of the instrument (Cronbach’s α=0.707). The “risk of isolation” was associated with a significant decrease in respondents’ tendency to stigmatize people with respiratory symptoms. However, when participants were concerned about “contagiousness of virus”, “danger of virus to their lives” or “lack of availability of protective equipment”, the stigma increased to a practical significance level.

**Table 2.** Typology of concern themes about COVID-19 with associated levels of psychological stress and severity of stigmatization of individuals with respiratory symptoms (effect size (SE) is weak at 0.2≤Cohen’s d≤0.49; p≤0.05)

| Table 2. Typology of concern themes about COVID-19 with associated levels of psychological stress and severity of stigmatization of individuals with respiratory symptoms (effect size (SE) is weak at $0.2 \leq \text{Cohen's } d \leq 0.49$; $p \leq 0.05$) | | | |
| --- | --- | --- | --- |
| Type of topics for anxieties | Prevalence<br>(persons / %) | Indicators change (SE): |  |
|  |  | stress | stigma |
| Risk to the lives and health of relatives | 1527 / 77,2 | – * | +0,06 |
| Possible financial difficulties | 1128 / 57,0 | +0,16 | -0,04 |
| Severe social consequences | 980 / 49,5 | +0,14 | -0,08 |
| Absence of specific treatment for COVID-19 | 789 / 39,9 | +0,1 | +0,19 |
| Impossibility of traditional way of life | 766 / 38,7 | +0,17 | -0,16 |
| Contagiousness of the virus | 708 / 35,8 | +0,1 | +0,27 |
| Fear for own life | 619 / 31,3 | +0,14 | +0,36 |
| Lack of protection equipment for sale | 544 / 27,5 | +0,16 | +0,23 |
| Possible lack of medication for daily intake | 434 / 21,9 | +0,39 | +0,19 |
| Risk of isolation | 351 / 17,7 | +0,43 | -0,14 |
| Footnote: *absence of statistically significant change in stress levels with this concern theme |  |  |  |

#### Psychological reactions of specific respondent groups

Among individual groups of respondents, specific concern themes had their own characteristics. The two concern themes most associated with psychological stress were identified in individuals who reported being diagnosed with affective disorders (Table 3). Moreover, the “risk of isolation” was more of a concern among those who had comorbid affective and somatic pathologies. However, the “possible lack of medication needed for daily intake” was more frequently the concern of respondents with affective disorders without somatic comorbidity.

**Table 3.** The specifics of anxiety experiences depending on the health group of respondents (effect size (SE) medium at 0.17≤Cramers’s V≤0.29)

| Table 3. The specifics of anxiety experiences depending on the health group of respondents (effect size (SE) medium at $0.17 \leq \text{Cramers's } V \leq 0.29$) | | | | | | |
| --- | --- | --- | --- | --- | --- | --- |
| Type of concern theme associated with COVID-19 |  | Healthy<br>N=643 | Presence of the disorder |  |  | Statistical significance criteria |
|  |  |  | affective<br>N=242 | somatic<br>N=737 | comorbid<br>N=336 |  |
| Risk of isolation | + | 11.0% | 21.5% | 17.1% | 31.0% | $\chi^2=63.8$; $p=0.000$<br>SE=0.25 |
|  | - | 89.0% | 78.5% | 82.1% | 69.0% |  |
| Lack of medication for daily intake | + | 22.2% | 34.3% | 14.4% | 30.4% | $\chi^2=59.6$; $p=0.000$<br>SE= 0.21 |
|  | - | 77.8% | 65.7% | 85.6% | 69.6% |  |

It is important to note that, among the 688 participants who had at least one of the two main concern themes associated with psychological stress, respondents without mental disorders were as common as those with affective disorders. An unexpected confirmation of the online questionnaire external validity was the identified prevalence of a specific concern for individuals with anxiety disorders about their life safety that distinguished them from respondents with mood disorders (Table 4).

**Table 4.** The specifics of anxiety experiences depending on type of affective disorder and gender (effect size (SE) medium at 0.1≤Cramers’s V≤0.3)

| Table 4. The specifics of anxiety experiences depending on type of affective disorder and gender (effect size (SE) medium at $0.1 \leq \text{Cramers's } V \leq 0.3$) | | | | |
| --- | --- | --- | --- | --- |
| Type of concern themes associated with COVID-19 |  | Representation by groups: |  | Statistical significance criteria |
|  |  | 1 | 2 |  |
| Gender: 1-male, 2-female |  |  |  |  |
| Lack of protection equipment for sale | + | 20.1% | 29.2% | $\chi^2=10.7$; p=0.001<br>SE=0.14 |
|  | - | 79.9% | 70.8% |  |
| Fear for own life | + | 23.1% | 33.2% | $\chi^2=12.4$; p=0.000<br>SE=0.14 |
|  | - | 76.9% | 66.8% |  |
| Type of affective disorder: 1- mood disorder, 2-anxiety disorder |  |  |  |  |
| Fear for own life | + | 26.7% | 40.7% | $\chi^2=8.8$; p=0.003 |
|  | - | 73.3% | 59.3% | SE=0.12 |

In addition to traditional populations considered most vulnerable to anxiety reactions (patients with affective and somatic disorders) many other cohorts have demonstrated various prevailing concerns about COVID-19. Women were more likely than men to worry about the absence of individual protective equipment in open sale and to fear for their own lives (Table 4). Single and unmarried participants, unemployed and employed in state institutions were more likely to fear isolation (Table 5).

**Table 5.** The specifics of anxiety experiences depending on occupational and family status (effect size (SE) medium at 0.15≤Cramers’s V≤0.25)

| Table 5. The specifics of anxiety experiences depending on occupational and family status (effect size (SE) medium at $0.15 \leq \text{Cramers's } V \leq 0.25$) | | | | | | | |
| --- | --- | --- | --- | --- | --- | --- | --- |
| Type of concern themes associated with COVID-19 |  | Representation by groups: |  |  |  |  | Statistical significance criteria |
|  |  | 1 | 2 | 3 | 4 | 5 |  |
| Occupation: 1-educational N = 271, 2-unemployed N = 435, 3-private hire N = 462, 4-state hiring N = 631, 5-own business N = 158 |  |  |  |  |  |  |  |
| Risk of isolation | + | 19.6% | 25.1% | 8.2% | 20.9% | 12.0% | $\chi^2=52.6$; $p=0.000$<br>SE=0.19 |
|  | - | 80.4% | 74.9% | 92.8% | 79.1% | 88.0% |  |
| Family: 1-widowers / widows N = 30, 2-divorced N = 144, 3-single N = 1014, 4-non-registered marriage N = 243, 5-registers marriage N = 526 |  |  |  |  |  |  |  |
| Risk of isolation | + | 3.3% | 13.2% | 22.5% | 16.9% | 11.8% | $\chi^2=34.5$; $p=0.000$<br>SE=0.16 |
|  | - | 96.7% | 86.8% | 77.5% | 83.1% | 88.2% |  |

Among respondents with higher education or academic degrees, as well as those over 31 years of age, there were significantly fewer concerns about the risk of isolation. A category of participants over 60 years of age tended to be more afraid of possible financial difficulties because of the pandemic (Table 6).

**Table 6.**
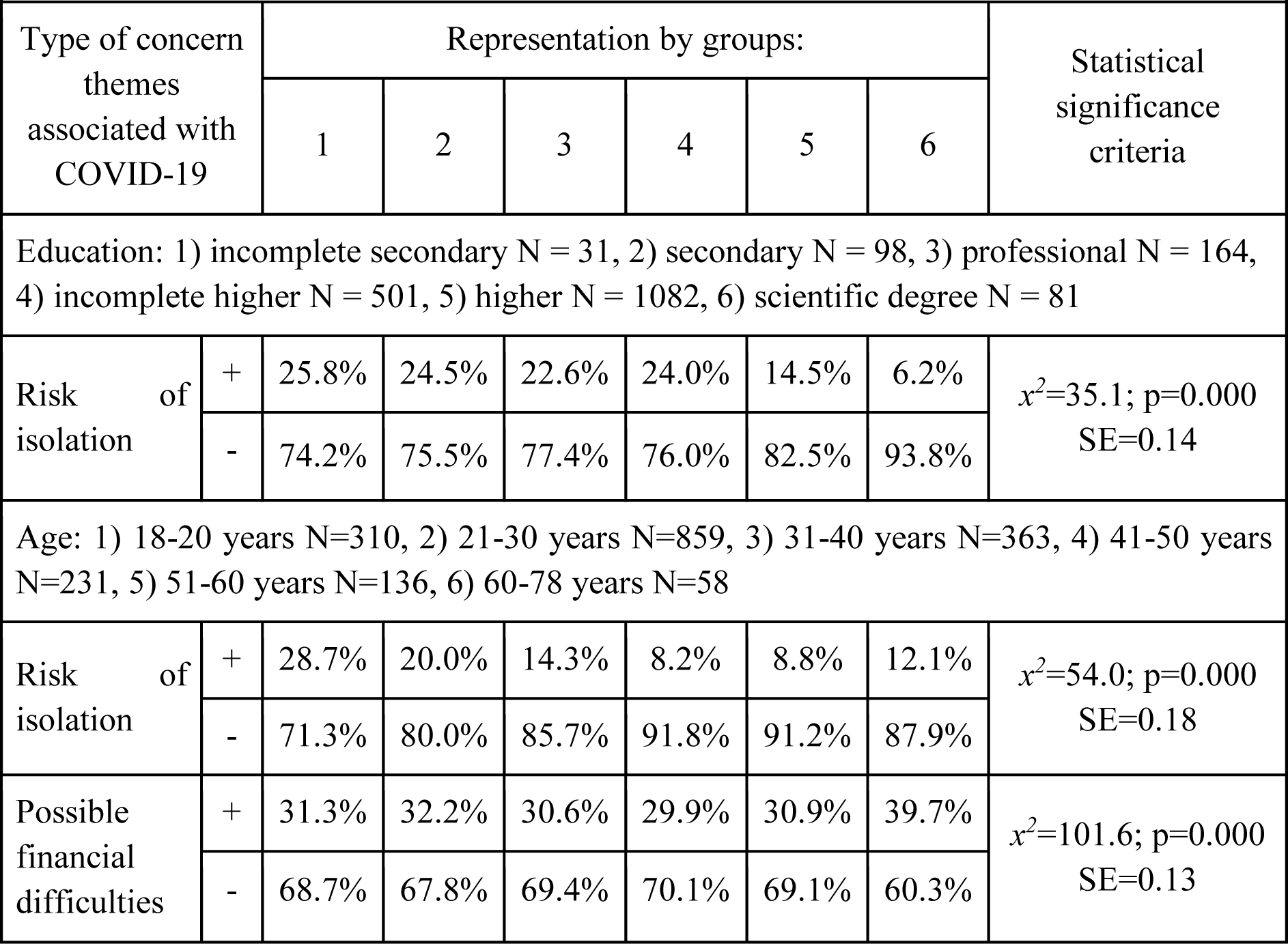
The specifics of anxiety experiences depending on educational level and age (effect size (SE) medium at 0.13≤Cramers’s V≤0.22)

## DISCUSSION

On the basis of an online survey, we obtained data that allow us to assess the structure of psychological experience specific for Russian-speaking respondents during the first week of the recommended self-isolation regime in Russia. The analysis showed high prevalence of variable anxiety trends associated with the COVID-19 pandemic among the study participants. Combined they increased the overall level of psychological stress in the evaluated sample.

Amidst the rapidly changing living conditions and activities associated with quarantine measures, respondents predictably presented with a variety of concerns related to the COVID-19 pandemic. It is essential to note that concerns about the “risk to the life and health of relatives and friends” did not lead to an increase in psychological stress. Because of that they can be considered within the framework of adaptive personality and psychological reactions. At the same time, the expansion of the concern themes led to the breakdown of adaptive mechanisms, provoking both the intensification of psychological and social stress. Increase in psychological stress manifested in higher anxiety, while social stress consciously or unconsciously was projected outside, causing increased stigma. It is important to emphasize that psychological stress escalated especially amid concerns about the “possible lack of medications for daily use” and the “risk of isolation”. In the first case such concerns are apparently associated with deterioration in subjective perception, and in the second case – with quarantine measures themselves provoking a wave of anxiety and anger. The increase in stigmatization attitudes turned out to be associated to the greatest extent with the following experiences: “fear for one’s own life”, “infectiousness of the virus”, and “lack off protective equipment sales”, which are, to a greater extent, caused by the feeling of loss of control over the situation.

Particular noteworthy is the data obtained from respondents who reported affective disorders diagnoses. For them, as well as for individuals who did not report any mental disorders, the same types of categories most associated with psychological stress were characteristic - “risk of isolation” and “inaccessibility of medications”. However, the “risk of isolation” was more concerning among those who had a somatic pathology combined with an affective one. At the same time, “the possible lack of medications necessary for daily use” often worried respondents suffering from affective disorders without somatic comorbidity. Moreover, people with anxiety disorders were characterized by the prevalence of a specific “fear for their own life”, in comparison with patients with affective disorders, which emphasizes the clinical diversity of their experiences.

The obtained data on the structure of the respondents’ anxious experiences make it possible to distinguish their characteristics among different population groups, which is important for further design of differentiated psychological and social assistance programs. In particular, “risk of isolation” concerns were most prevalent in young respondents (under 31 years old) who were single / unmarried, had no higher education and were unemployed and also in individuals with a combined affective and somatic pathology. If in the first three cases this may be due to personal immaturity, unformed skills of self-control and self-employment, as well as a temporary loss of the ability to communicate. For the unemployed the main reason is a decrease in the possibility of material support. For people of the older age group, “material difficulties” turned out to be a special topic of anxiety, which obviously dictates the need for other informational and social interventions.

The WHO COVID-19 strategic preparedness and response plan has not yet identified any strategies to address emerging mental health needs [13], although the need for them will most likely increase both during the epidemic and after it.

The scientific literature available to us does not contain data on psychological reactions at the initial stages of the epidemiological situation deterioration and the announcement of quarantine measures, as a fact of official recognition of the increase in epidemiological distress. In China, which was the first to deal with the organization on all levels of medical care to limit spread of coronavirus, the Principle for Supporting Mental Well-being was developed. It included: 1) determining the current status of mental health in the population, 2) determining the circle of people with a high risk of suicide and aggression, 3) developing structured measures of care [14]. But the effectiveness of the psychological assistance in this region was considered insufficient, which was associated with a lack of experience in teaching the principles of maintaining mental health [15].

Thus, the size and social heterogeneity of the risk group necessitate the use of broad social interventions to overcome the socio-psychological consequences of the pandemic. They can be implemented according to the principle of aid separation and should include: the stage of psycho-social support, special psychological assistance, as well as clinical and psychological assistance involving psychiatrists. Based on the experience of organizing such work in China and also considering the data we have received, one of the obstacles to organizing effective support for the population can be stigma / discrimination [16].

## CONCLUSIONS

Intervention in a psychological crisis should be considered as an important part of a public health response to the outbreak of COVID-19. The first step should be the widespread involvement of competent professionals in the area of infectious diseases, epidemiology and mental health in order to inform citizens about effective measures to limit the further spread of infection and ways to prevent the increase in stress in the media, as opposed to amateurish judgments that flood the Internet. Furthermore, our data allow us to recommend the correction of distress concerns broadly, but also with more precision. Predominant attention should be paid not only to patients with affective disorders, but also to young people; women; individuals without higher education, not employed in labor or educational activities, single and unmarried. The most urgent need for correction of anxiety about the COVID-19 pandemic was identified in the concerns about the possible lack of medications for daily use and the risk of isolation, since these types of experiences are associated with the maximum increase in anxiety distress in the evaluated sample. It is necessary to create access to individual psychological counseling on-line at the state level and provide psychological and psychiatric assistance to those in need, while ensuring adequate epidemiological safety. The timely provision of psychological and psychiatric care to the ones in need is especially important, since psychological stability also contributes to the preservation of the physical health of the population.

## Data Availability

The data that support the findings of this study are available from the corresponding author upon reasonable request.

## Acknowledgements

We want to thank The Russian Society of Psychiatrists (RSP), Anastasia Petrova and ANPO “Partnership of Equal”, Maria Pushkina (Favorskaya) and Association “Bipolar”, Victor Lebedev and project “Pinel’s affair”, and educational portal “Psychiatry & Neurosciences”.

## Notes

**Conflict of interests:** None to declare.

### Competing Interest Statement

The authors have declared no competing interest.

### Funding Statement

none

